# Deep learning-based model for detecting 2019 novel coronavirus pneumonia on high-resolution computed tomography: a prospective study

**DOI:** 10.1101/2020.02.25.20021568

**Authors:** Jun Chen, Lianlian Wu, Jun Zhang, Liang Zhang, Dexin Gong, Yilin Zhao, Shan Hu, Yonggui Wang, Xiao Hu, Biqing Zheng, Kuo Zhang, Huiling Wu, Zehua Dong, Youming Xu, Yijie Zhu, Xi Chen, Lilei Yu, Honggang Yu

## Abstract

**Background:** Computed tomography (CT) is the preferred imaging method for diagnosing 2019 novel coronavirus (COVID19) pneumonia. Our research aimed to construct a system based on deep learning for detecting COVID-19 pneumonia on high resolution CT, relieve working pressure of radiologists and contribute to the control of the epidemic.

**Methods:** For model development and validation, 46,096 anonymous images from 106 admitted patients, including 51 patients of laboratory confirmed COVID-19 pneumonia and 55 control patients of other diseases in Renmin Hospital of Wuhan University (Wuhan, Hubei province, China) were retrospectively collected and processed. Twenty-seven consecutive patients undergoing CT scans in Feb, 5, 2020 in Renmin Hospital of Wuhan University were prospectively collected to evaluate and compare the efficiency of radiologists against 2019-CoV pneumonia with that of the model.

**Findings:** The model achieved a per-patient sensitivity of 100%, specificity of 93.55%, accuracy of 95.24%, PPV of 84.62%, and NPV of 100%; a per-image sensitivity of 94.34%, specificity of 99.16%, accuracy of 98.85%, PPV of 88.37%, and NPV of 99.61% in retrospective dataset. For 27 prospective patients, the model achieved a comparable performance to that of expert radiologist. With the assistance of the model, the reading time of radiologists was greatly decreased by 65%.

**Conclusion:** The deep learning model showed a comparable performance with expert radiologist, and greatly improve the efficiency of radiologists in clinical practice. It holds great potential to relieve the pressure of frontline radiologists, improve early diagnosis, isolation and treatment, and thus contribute to the control of the epidemic.

## Introduction

In December 2019, a new coronavirus infection disease (hereinafter referred to as COVID-19) was first reported in Wuhan. Subsequently, the outbreak began to spread widely in China and even abroad.[1-3]

The clinical manifestations of the COVID-19 pneumonia is complicated and could be characterized as fever, cough, myalgia, headache, and gastrointestinal symptoms onset.[4] Although the nucleic acid detection was considered determinant for identifying the COVID-19 infection and more rapid detection kit for the novel coronavirus has come into mass production, computed tomography (CT) scan is still the most efficient modality for detecting and evaluating the severity of pneumonia.[5] An update series demonstrate that CT findings were positive in all 140 laboratory-confirmed COVID-19 patients, even in the early stage.[4,6] In the fifth version of diagnostic manual of COVID-19 launched by the National Health and Health Commission of China, the radiographic characteristics of pneumonia was included the clinical diagnostic standard in Hubei Province.[7] Subsequently, 14,840 new cases of COVID-19 were reported within one day on Feb 13, 2020 in Wuhan, including 13332 cases of clinical diagnoses.[8] This highlighted the importance of CT in the diagnosis of COVID-19 pneumonia.

Due to the outbreak of the COVID-19, thousands of patients waited in line for CT examination in the designated fever outpatient hospital at Wuhan and other cities. As of Feb 14, there are 5,534 suspected cases, 38,107 confirmed patients receiving treatment in hospital, and 77,323 cases under medical observation in Hubei province.[9] Most of them need to undergo CT examination, however, there are less than 4,500 radiologists in cities of Hubei according to the China Health Statistical Yearbook (2018).[10] Meanwhile, because the lung infection foci are small in the early stage of the COVID-19 infection, thinner layer (2.5mm, 1.25mm or even 0.625mm) scanning were usually needed instead of conventional CT scan (5 mm) for diagnosis, which would be more time-consuming. All these made radiologists overloaded, delay the diagnosis and isolation of patients, affect patient’s treatment and prognosis, and ultimately, affect the control of COVID-19 epidemic.

Deep learning, an important breakthrough in the domain of AI in the past decade, has huge potential at extracting tiny features in image analysis.[11] Our group also succeeded in recruiting this technique in minor lesion detection and real-time assistance to doctors in gastrointestinal endoscopy.[12-16]

In the present research, we construct and validate a system based on deep learning for identification of viral pneumonia on CT. Our model has comparable performance with expert radiologist, but take much less time.

## Method

### Patients

We first retrospectively collected 46,096 anonymous images from 106 admitted patients, including 51 patients of laboratory confirmed COVID-19 pneumonia and 55 control patients of other diseases in Renmin Hospital of Wuhan University (Wuhan, Hubei province, China) for model development. The patients’ CT scan images and reports, history, clinical manifestations, physical findings, and viral pathogen results were all collected. For prospective patients, 27 consecutive patients undergoing CT scans were enrolled in the designated CT rooms in Feb 5, 2020 in Renmin Hospital of Wuhan University.

This study was approved by the Ethics Committee of Renmin Hospital of Wuhan University. Written informed consent was provided by all prospective participants. Because of virus contamination, the signed informed consents were carefully sealed and kept in the specific place according to the Law of the People’s Republic of China on Infectious Disease Prevention and Control.[17] For patients whose CT scans were stored in the retrospective databases, informed consent was waived by the Ethics Committee.

### Diagnostic testing for COVID-19

Patient’s respiratory secretions were collected and transferred to a sterile test tube with a virus transport medium. Fluorescent RT-PCR analysis of samples was performed using the COVID-19 nucleic acid detection kit developed by Shanghai Geneodx Biotechnology Co., Ltd. This detection kit was approved by the US National Drug Administration (NMPA) on January 26, 2019 and recommended by the Centers for Disease Control and Prevention (CDC).[18] The rapid, high-precision COVID-19 detection kit greatly accelerated the confirmation of human COVID-19 infection.

### Datasets

As shown in Figure 1, a total of 46,096 CT scan images from 51 COVID-19 pneumonia patients and 55 control patients of other disease were collected for developing the model to detect COVID-19 pneumonia. After filtering those images without good lung fields, 35355 images were selected and split into training and retrospectively testing datasets. Enrolled images in training dataset covered almost all common CT features of COVID19 pneumonia, as presented in Figure 2. Three radiologists with more than 5 years of clinical experience labelled infection lesions of COVID-19 pneumonia patients in training dataset, and selected images containing COVID19 pneumonia lesions in testing set, and their labels were combined by consensus. For prospectively testing the model, 13,911 images of 27 consecutive patients undergoing CT scans in Feb 5, 2020 in Renmin Hospital of Wuhan University were further collected. All CT scans were obtained in Renmin Hospital of Wuhan University. The instruments used in this study included Optima CT680, Revolution CT and Bright Speed CT scanner (all GE Healthcare).

**Figure 1.**
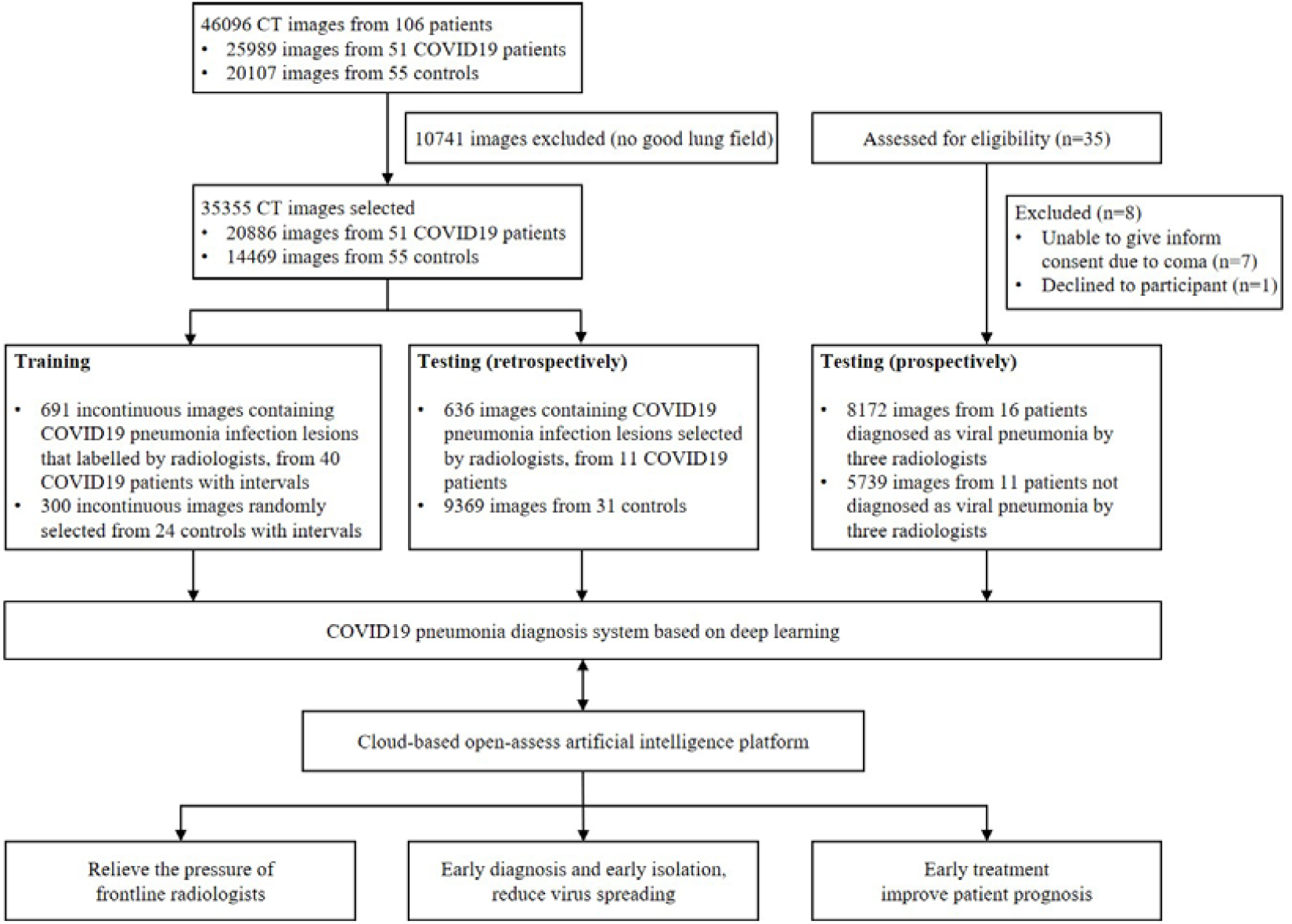
Workflow diagram for the development and evaluation of the model for detecting COVID19 pneumonia.

**Figure 2.**
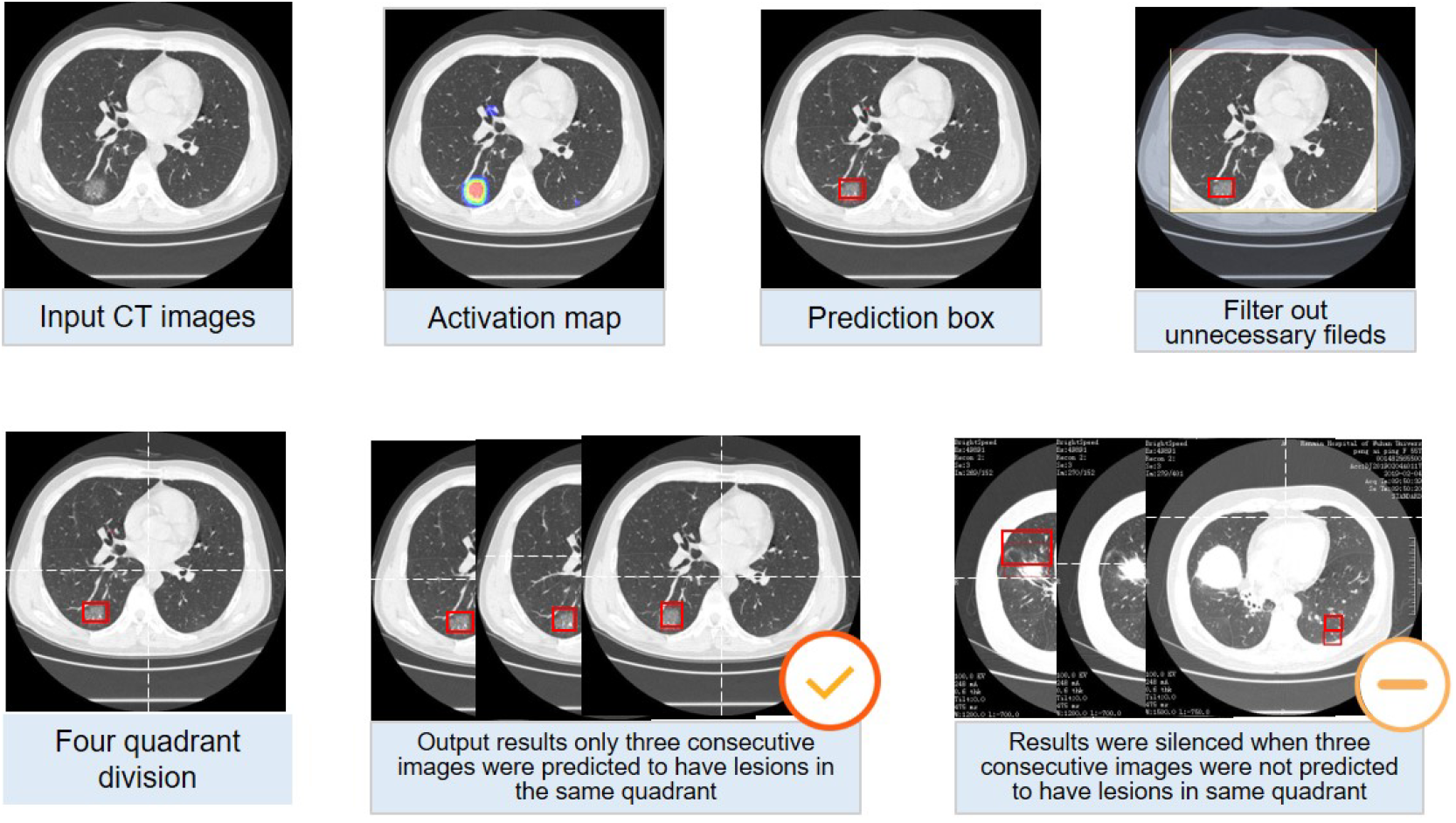
Representative images of COVID19 pneumonia. More than six common Computed tomography (CT) features of COVID19 pneumonia were covered in selected images. 1a-1d, the lesions were mainly ground-glass-like, with thickened blood vessels walking and including gas-bronchial signs in 1c; 2a-2d, the lesions were mainly ground glass changes, and paving stone-like changes were observed on 2d; 3a-3c, the lesions become solid with a large range, and air-bronchial signs are seen inside; 4, the lesion is located in the lower lobe of both lungs, and is mainly grid-like change with ground glass lesion; 5a-5b, the lesions are mainly consolidation; 6a-6b, the lesions are mainly large ground glass shadows, showing white lung-like changes, with air-bronchial signs.

### Training algorithm

UNet++, a novel and powerful architecture for medical image segmentation was implemented to develop the model.[19,20] We first trained UNet++ to extract valid areas in CT images using 289 randomly selected CT images and tested it in other 600 randomly selected CT images. The training images were labelled with the smallest rectangle containing all valid areas by researchers. The model successfully extracted valid areas in 600 images in testing set with an accuracy of 100%. For detecting suspicious lesions on CT scans, 691 images of COVID-19 pneumonia infection lesions labelled by radiologists and 300 images randomly selected from patients of non-COVID-19 pneumonia were used. Taking the raw CT scan images as input with a resolution of 512×512, and the labelled map from the expert as output, UNet++ was used to train in Keras in an image-to-image manner. The suspicious region was predicted under a confidence cutoff value of 0.50, and a prediction box pixel of over 25. The training curves of UNet++ for extracting valid areas and detecting suspicious lesions in CT images were shown in **Supplementary Figure 1** and **Supplementary Figure 2**, respectively. The prediction schematic of the model was shown in Figure 3. Raw images were firstly input into the model, and after processing of the model, prediction boxes framing suspicious lesions were output. Valid areas were further extracted and unnecessary fields were filter out to avoid possible false positives. To predict by case, a logic linking the prediction results of consecutive images was added. CT images with the above prediction results were divided into four quadrants, and results would be output only when three consecutive images were predicted to have lesions in the same quadrant.

**Figure 3.**
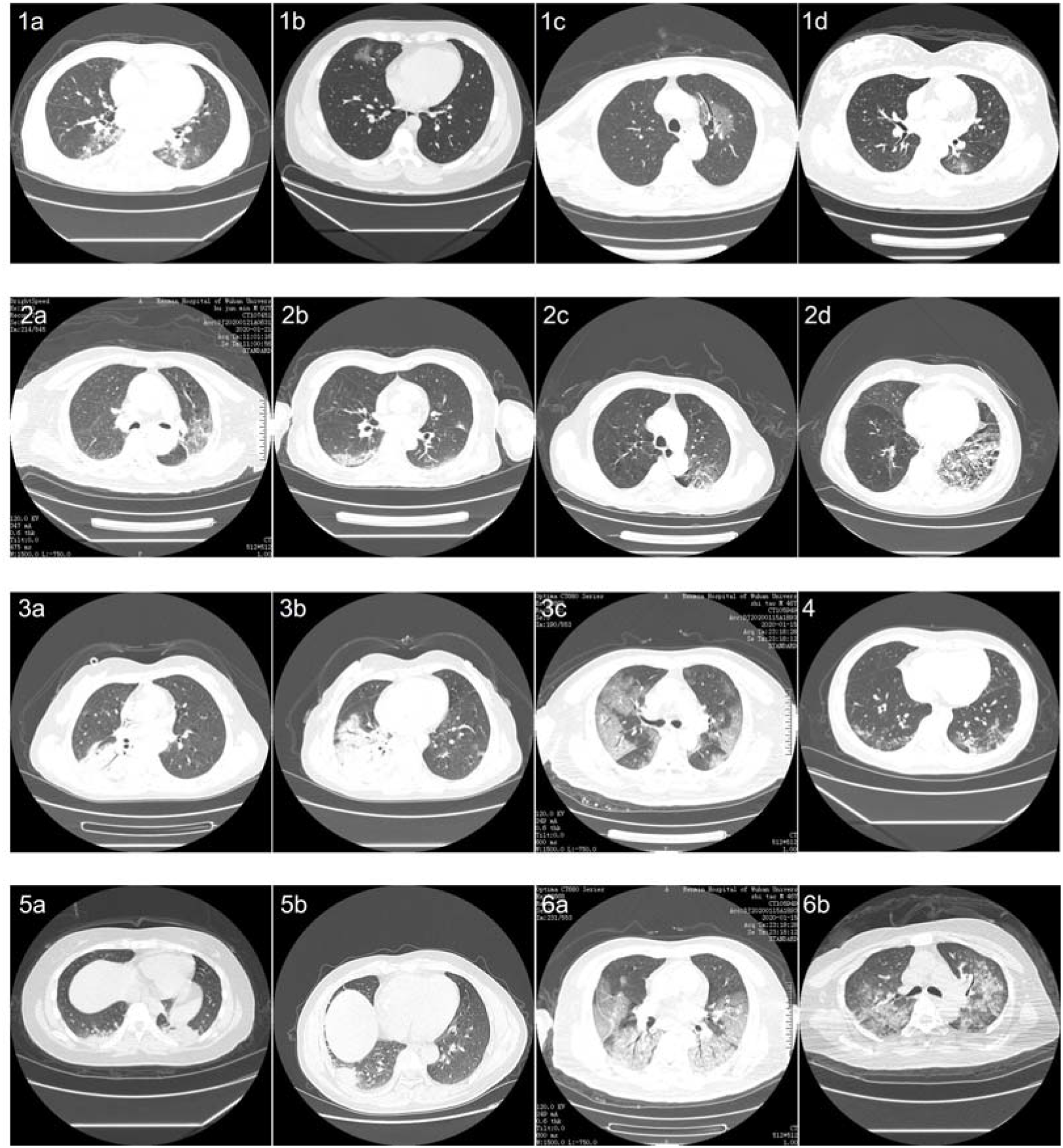
Processing and prediction schematic of the model. Raw images were firstly input into the model, and after processing of the model, prediction boxes framing suspicious lesions were output. Valid areas were further extracted and unnecessary fields were filter out to avoid possible false positives. To predict by case, a logic linking the prediction results of consecutive images was added. Computed tomography (CT) images with the above prediction results were divided into four quadrants, and results would be output only when three consecutive images were predicted to have lesions in the same quadrant.

### Testing of the model in retrospective data

To evaluate the performance of the model on CT scan images, five metrics including the accuracy, sensitivity, specificity, PPV and NPV were calculated as follows: accuracy = true predictions/total number of cases, sensitivity = true positive/positive, specificity = true negative/negative, positive prediction value (PPV) = true positive/(true positive + false positive), negative prediction value (NPV) = true negative/(true negative + false negative). The “true positive” is the number of correctly predicted COVID-19 pneumonia cases/images, “false positive” is the number of mistakenly predicted COVID-19 pneumonia cases/images, “positive” is the number of cases/images of COVID-19 pneumonia patients, “true negative” is the number of correctly predicted non-COVID-19 pneumonia cases/images, “false negative” is the number of mistakenly predicted non-COVID-19 pneumonia cases/images and ‘negative’ is the number of non-COVID-19 pneumonia cases/images enrolled. For image-based metrics, 636 images containing infection lesions identified by radiologists among 11 patients of COVID-19 pneumonia were used as the positive sample, and 9369 CT scan images from 31 patients of non-COVID-19 pneumonia were used as the negative sample.

### Evaluating the efficiency of radiologist in traditional way

To evaluate the performance and cost of time of radiologist against 2019-CoV pneumonia, prospectively consecutive patients undergoing CT scans were enrolled in the designated CT rooms in Feb 5, 2020 in Renmin Hospital of Wuhan University. An expert radiologist was required to read all CT images of enrolled patients using the working computer, and determine if each patient has viral pneumonia. The research assistant used a stopwatch to record the expert’s reading time. The expert radiologist was associate chief physician of the Radiology Department of Renmin Hospital of Wuhan University, with clinical experience of 30 years, and independently diagnosed about 300 viral pneumonia. Hospitalized viral pneumonia cases judged by radiologists were all diagnosed using COVID-19 nucleic acid detection kit to confirm COVID-19 infection. The computed radiography imaging system used by the radiologist was VisionPACS (Intechhosun, Being, China).

### Comparison between the model and radiologist in prospective data

The CT scan images of the prospective patients as above were collected and imported into the model for prediction. The model’s performance and cost of time were compared with that of the expert radiologist. Inconsistent results between the expert and model were reviewed by three radiologists, including the expert and other two radiologists, senior staff members of the Radiology Department of Renmin Hospital of Wuhan University, with clinical experience about 10 years, and independently diagnosed about 150 viral pneumonia.

### Evaluating the efficiency of radiologist with the assistance of AI

To evaluate the performance and cost of time of radiologist against 2019-CoV pneumonia with the assistance of our model, the prediction results of the model (whether a patient has viral pneumonia, and labels marking lesions) were copied to the working computer in the designated CT rooms. After 10 days of wash out period (in Feb 16, 2020), the same expert radiologist was required to re-read all CT images of 27 prospective patients using the working computer where results of the model could be viewed, and determine if each patient has viral pneumonia. The research assistant used a stopwatch to record the expert’s reading time again. Hospitalized viral pneumonia cases judged by radiologists were all diagnosed using COVID-19 nucleic acid detection kit to confirm COVID-19 infection. The computed radiography imaging system used by the radiologist was VisionPACS (Intechhosun, Being, China).

### Statistical analysis

A two-tailed paired Student’s t test with a significance level of 0.05 was used to compare differences in the cost time of the model and radiologist.

## RESULTS

### Patients

The baseline characteristics and CT findings of 51 patients of 2019-CoV pneumonia and 55 control patients in retrospective dataset were shown in Table 1 and Table 2, respectively. Baseline characteristics were comparable between training and testing datasets. The 31 control patients in retrospective testing dataset include 2 lung cancer, 4 tuberculosis, 2 bronchiectasis, 2 nonviral pneumonia, 1 lung bullae and 20 with no obvious finding in CT scan.

**Table 1.** Clinical characteristics of enrolled patients of COVID19 pneumonia

|  | All patients (n=51) | Training set (n=40) | Testing set (n=11) |
| --- | --- | --- | --- |
| Age, years, median (IQR) | 52 (38, 69) | 54.5 (41.5, 71.25) | 42 (34.5, 65.5) |
| Sex, n (%) |  |  |  |
| Men | 18 (35.3) | 11 (27.5) | 7 (63.6) |
| Women | 33 (64.7) | 29 (72.5) | 4 (36.4) |
| Presenting symptoms and signs onset, n (%) |  |  |  |
| fever | 28 (54.9) | 21 (52.5) | 7 (63.6) |
| cough | 27 (52.9) | 20 (50) | 7 (63.6) |
| Chest tightness or pain | 7 (13.7) | 6 (15) | 1 (9.1) |
| dyspnea | 6 (11.8) | 6 (15) | 0 |
| Muscle soreness | 10 (19.6) | 8 (20) | 2 (18.2) |
| Expectoration | 12 (23.5) | 10 (25) | 2 (18.2) |
| Headache | 3 (5.9) | 2 (5) | 1 (9.1) |
| Digestive symptoms | 6 (11.8) | 6 (15) | 0 |
| CT findings, n (%) |  |  |  |
| Unilateral pneumonia | 18 (35.3) | 13 (30) | 5 (45.5) |
| bilateral pneumonia | 33 (64.7) | 27 (70) | 6 (54.5) |
| Multiple mottling and ground-glass opacity | 16 (31.4) | 13 (32.5) | 3 (27.3) |

**Table 2.** Clinical characteristics of enrolled control patients.

|  | All patients (n=55) | Training set (n=24) | Testing set (n=31) |
| --- | --- | --- | --- |
| Age, years, median (IQR) | 48(34.5, 55) | 50.5 (38.25, 55.25) | 47 (34.5, 54.5) |
| Sex, n (%) |  |  |  |
| Men | 31 ( 56.36 ) | 12 (50) | 19 (61.29) |
| Women | 24 ( 43.64 ) | 12 (50) | 12 (38.71) |
| CT examination indications, n (%) |  |  |  |
| Viral pneumonia to be discharged | 7 ( 12.73 ) | 0 ( 0 ) | 1 ( 3.23 ) |
| Pulmonary bullae to be discharged | 2 ( 3.64 ) | 2 ( 8.33 ) | 0 ( 0 ) |
| Tuberculosis | 1 ( 1.82 ) | 1 ( 4.17 ) | 0 ( 0 ) |
| Lower respiratory infection | 3 ( 5.45 ) | 1 ( 4.17 ) | 2 ( 6.45 ) |
| Metastatic lung tumor to be discharged | 7 ( 12.7 ) | 5 ( 20.83 ) | 2 ( 6.45 ) |
| Routine examination before admission | 41 ( 74.55 ) | 15 ( 62.5 ) | 26 ( 83.87 ) |
| CT findings, n (%) |  |  |  |
| No obvious abnormality | 44 ( 80 ) | 24 | 20 ( 64.52 ) |
| Tuberculous lesion | 4 ( 7.27 ) | -- | 4 ( 12.90 ) |
| Suspected neoplastic lesions | 2 ( 3.64 ) | -- | 2 ( 6.45 ) |
| Inflammatory lesions (non-viral) | 2 ( 3.64 ) | -- | 2 ( 6.45 ) |
| Bronchiectasia | 2 ( 3.64 ) | -- | 2 ( 6.45 ) |
| Bullae of lung | 1 ( 1.82 ) | -- | 1 ( 3.23 ) |

### The performance of the model on retrospective dataset

Among 4382 CT images from 11 patients of COVID-19 pneumonia and 9369 images from 31 control patients, the model correctly diagnosed the patients with a per-patient sensitivity of 100%, specificity of 93.55%, accuracy of 95.24%, PPV of 84.62%, and NPV of 100%. A per-image sensitivity of 94.34%, specificity of 99.16%, accuracy of 98.85%, PPV of 88.37%, and NPV of 99.61%. Representative images predicted by the model were shown in Figure 4.

**Figure 4.**
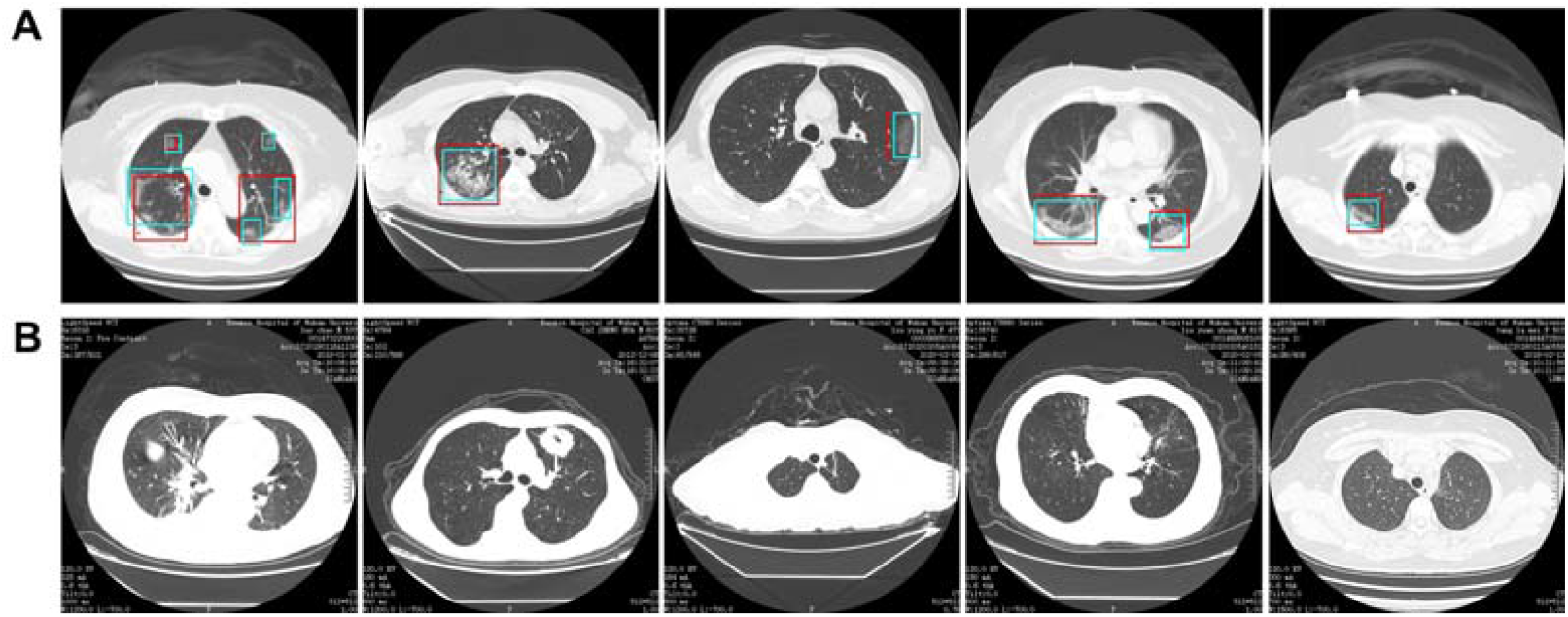
Representative images of the model’s predictions. A. Computed tomography (CT) images of COVID19 pneumonia. The predictions between the artificial intelligence model and radiologists were consistent. Green boxes, labels from radiologists; red boxes, labels from the model. B. CT images of the control. The first image is an ordinary bacterial pneumonia, showing a consolidation of the right lower lobe. The second image has a tumorous lesion in the lung, showing a mass in the left upper lobe, with burrs seen at the edges, and showing leaf-like growth with vacuoles inside. The third image is a secondary pulmonary tuberculosis, showing a left apical fibrous cord. The fourth image is a bronchiectasis complicated with infection, showing bronchodilation and expansion, cystic changes, and surrounding patches of infection. The fifth image shows normal lungs.

### The performance of the model in consecutive prospective patients

Twenty-seven patients were enrolled in the prospective dataset. Sixteen (59.26%) patients were diagnosed as viral pneumonia by the expert radiologist, and the other eleven patients were not. Two other radiologists reviewed the CT imaging, approved the expert’s results, and summarized that the CT characteristics of the 11 patients not diagnosed by the expert include 5 ground glass nodules, 3 diminutive nodules, 2 normal and 1 fibrosclerosis.

The model successfully detected all the 16 patients of viral pneumonia diagnosed by the expert. Among the other 11 patients, 2 were also detected by the model. The predictions in one case was fibrosclerosis lesion, and the other one was normal stomach bubble. Using results of the radiologists as the gold standard, the model achieved a per-patient sensitivity of 100%, accuracy of 92.59%, specificity of 81.82%, PPV of 88.89% and NPV of 100% in the 27 prospective patients. Among the 16 patients diagnosed as viral pneumonia by radiologists, 8 admitted patients were confirmed as COVID-19 infection, and the others were outpatients that difficult to follow nucleic acid results. The average prediction time for model was 41.34s per patient (IQR 39.76-44.48). The performance of the model on detecting COVID-19 pneumonia was shown in Table 3.

**Table 3.** The performance of the deep learning model on both retrospective and prospective dataset

|  | Sensitivity | Specificity | Accuracy | PPV | NPV |
| --- | --- | --- | --- | --- | --- |
| Retrospective testing |  |  |  |  |  |
| Per patient | 100% | 93.55% | 95.24% | 84.62% | 100% |
| Per image | 94.34% | 99.16% | 98.85% | 88.37% | 99.61% |
| Prospective testing<br>(Per patient) | 100% | 81.82% | 92.59% | 88.89% | 100% |

\*PPV: positive prediction value, NPV, negative prediction value.

### Comparison between the efficiency of radiologist with or without the assistance of AI

In the first time the expert radiologist read CT scan images of the 27 prospective patients, the average reading time for him to determine whether each patient has viral pneumonia was 116.12s per case (IQR 85.69-118.17). After 10 days of wash out period, the same expert radiologist re-read the CT images of the 27 prospective patients with the assistance of the AI model. The results for determining whether each patient has viral pneumonia were not changed, while the average reading time of the expert was greatly decreased by 65%. This indicates that the efficiency of radiologist could be greatly improved with the assistance of AI.

A website has been made available to provide free access to the present model (http://121.40.75.149/znyx-ncov/index) (Figure 5). CT scan images could be uploaded by both clinicians and researches as a second opinion consulting service, especially in other provinces or countries unfamiliar with the radiologic characteristics of COVID-19. Cases of COVID-19 pneumonia were also been made available on the open-access website, which might be a useful resource for radiologists and researchers for fighting COVID-19 pneumonia.

**Figure 5.**
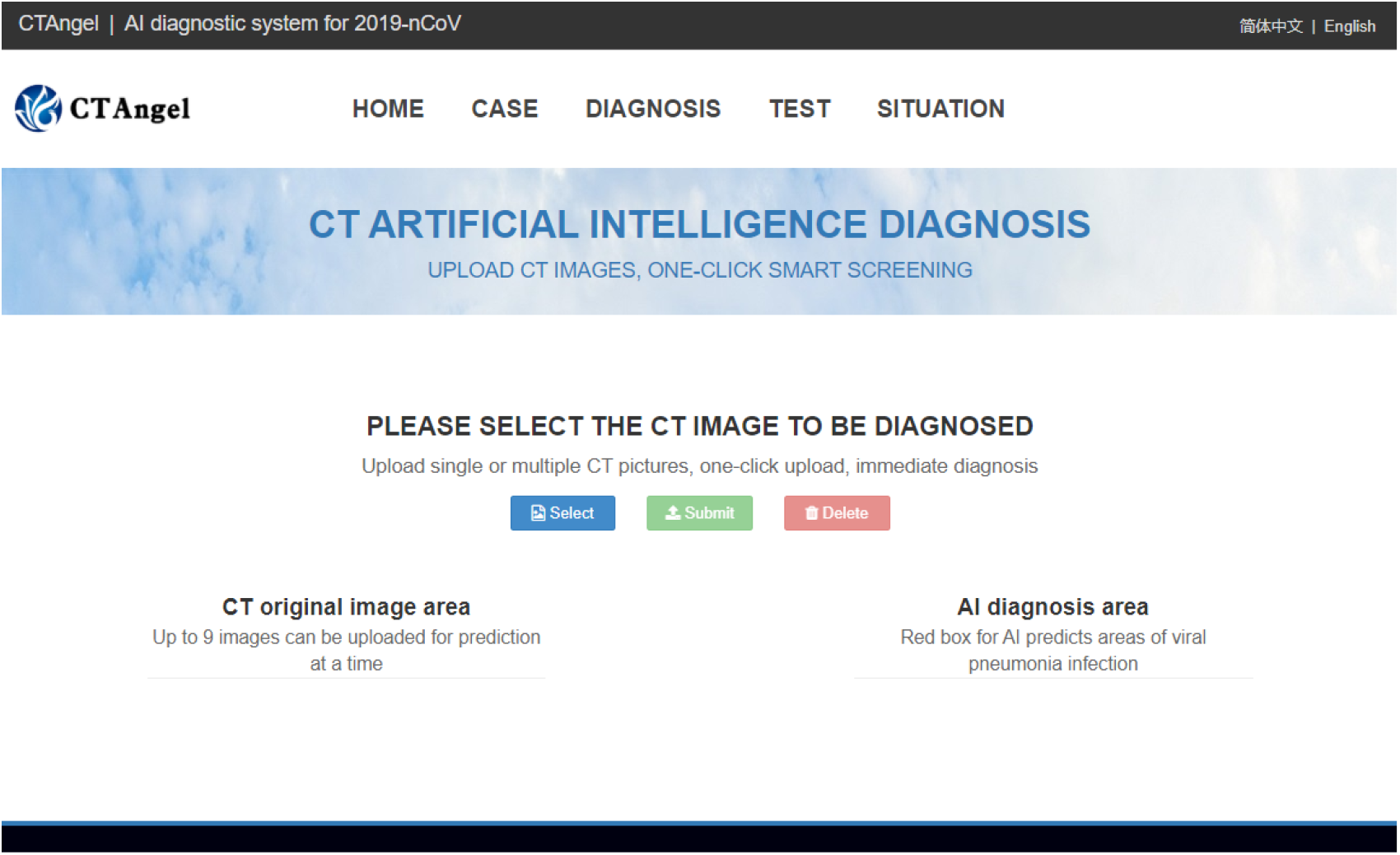
Main interface of the open-access artificial intelligence platform which provides fast and sensitive assistance for detecting COVID19 pneumonia. (http://121.40.75.149/znyx-ncov/index)

## DISCUSSION

As of Feb 14, 2020, the national health commission had reported 66,492 confirmed cases, 1,523 deaths and 8,969 suspected cases.[21] In the face of such large number of patients and high contagiosity of the novel coronavirus (with an estimated reproduction number R0 of 2.2∼6.47), timely diagnosis and isolation are the keys to prevent further spread of the virus.[22-26] CT scan is the most efficient modality for screening and clinically diagnosing COVID-19 pneumonia.[5,7] However, compared to the needs of the patients, the number of radiologists is quite small, especially in Hubei province, China, which could greatly delay the diagnosis and isolation of patients, affect patient’s treatment and prognosis, and ultimately, affect the overall control of COVID-19 epidemic.

Deep learning, a technology has shown great performance on extracting tiny features in radiology data, may hold the promise to alleviate this problem.[11] Recently, Ardila D, et al achieved end-to-end lung cancer screening on low-dose chest CT with an AUC of 94.4%.[27] Chae KJ, et al successfully used the convolutional neural network to classify small (≤2 cm) pulmonary nodules on CT scan images.[28] However, there was rare research being conducted to detect viral pneumonia.[11,27,28] In previous work, our group succeeded in recruiting deep learning in minor lesion detection and real-time assistance to doctors in gastrointestinal endoscopy.[12-16] Here, we enrolled this technique in identification of COVID-19 pneumonia in CT images. Results from both retrospective and prospective patients showed that the model was comparable to the level of expert radiologist, and hold great potential to reduce diagnosing time. (Figure 6)

**Figure 6.**
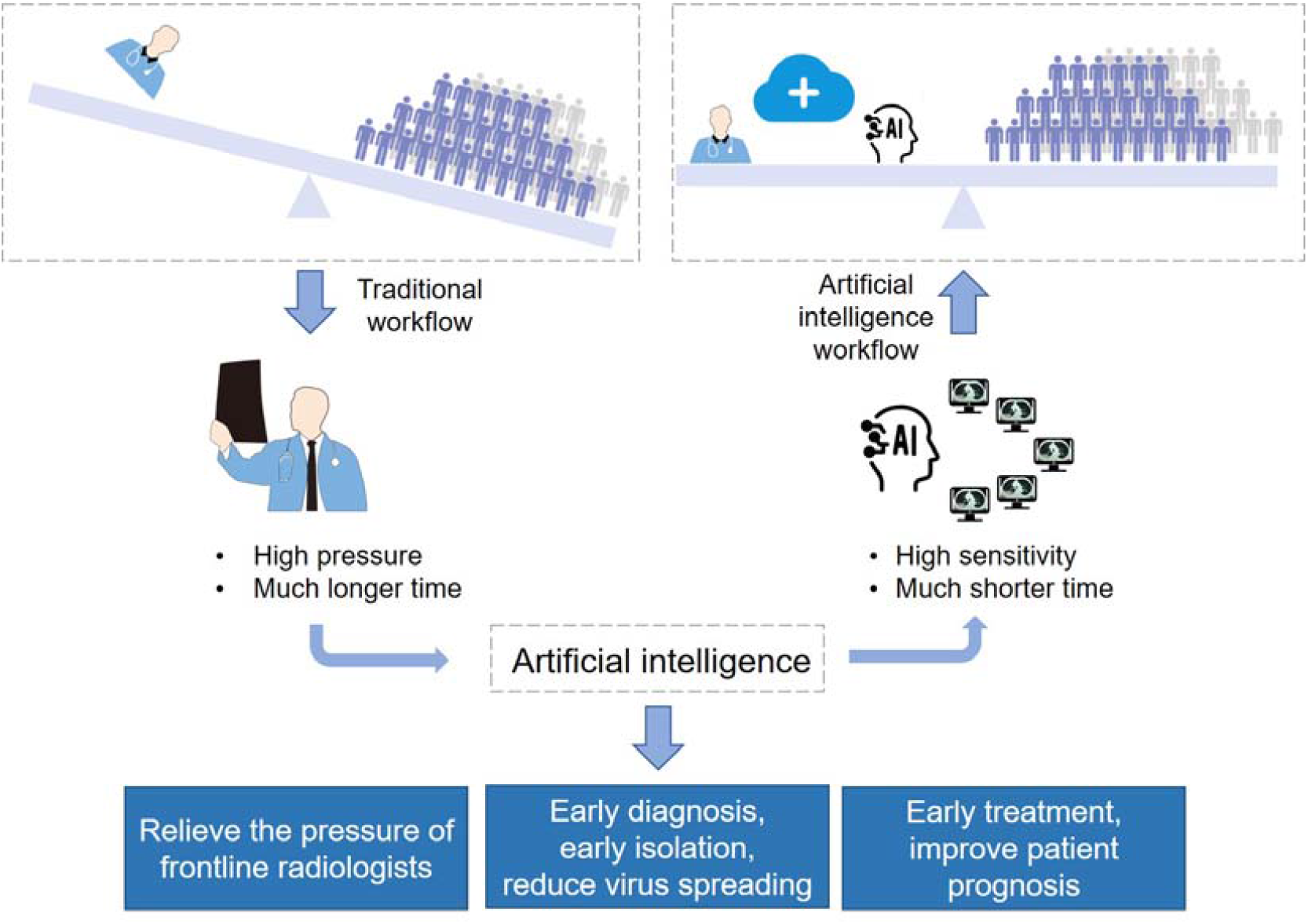
Abstract diagram. Computed tomography (CT) is the most efficient modality for screening and clinically diagnosing COVID-19 pneumonia. However, compared to the needs of the patients, the number of radiologists is quite small. After enrolling artificial intelligence in identifying COVID-19 pneumonia in CT images, the efficiency of diagnosis is greatly improved. The artificial intelligence holds great potential to relieve the pressure of frontline radiologists, accelerates the diagnosis, isolation and treatment of COVID19 patients, and therefore contribute to the control of the epidemic.

Early diagnosis and early isolation of suspected patients are the most important ways to prevent the spread of epidemic.^19^ Due to the sudden outbreak of COVID19, the radiology department is overloaded and patients have to wait for long times for chest CT scan, which largely increase the risk of cross-infection. In recent days, radiologists’ daily workload is huge in Hubei province, and a CT scan report has to be awaited several hours to achieve. Based on the number of suspected patients and close contacts in being, radiologists in the hardest hit, Hubei province, China, may not be enough to resist the rapid spread of the virus, which holds high estimated R0 of 2.2∼6.47.[23-26] It could be inferred that before radiologists fulfilling the demands of existing patients, newly infected cases would appear, and the overall burden of radiologists is more overwhelming like a growing snowball. Relieving the pressure of radiologists is essential for the control of virus spreading. In the present study, our model achieved a comparable performance but with much shorter time compared with expert radiologists. It holds great potential to relieve the pressure of radiologists in clinical practice, and contribute to the control of the epidemic.

Timely diagnosis and early treatment of infected patients is important for patients’ prognosis.[29] The fatality rate of COVID19 patients in Hubei province is significantly higher than that of other regions, which probably due to delayed treatment and shortage of medical resources.[8,30] Accelerating diagnosis efficiency is significant for improving patient outcomes. In the present study, our model helped expert radiologists achieve the same work with much shorter time, which greatly accelerats the efficiency of diagnosis in clinical practice, and may contribute to the improvement of patient outcome.

In addition to relieving radiologists’ pressure and accelerating diagnosis efficiency, artificial intelligence also holds the potential to reduce miss diagnosis of COVID-19 patients. The lung infection foci are sometimes mild in the early stage of the COVID-19 infection,^5^ and requires careful observation under 0.625mm layer scanning. Radiologists vary in skills, and could be affected by subjective status and outside pressure. One miss diagnosis could lead to multiple spread. The model is highly sensitive and stable, and would never be affected by work burden and work time. As a preliminary screening tool, it might help radiologists improve the sensitivity and reduce miss diagnosis.

On the basis of the accuracy and efficiency of the model in detecting COVID-19 pneumonia, a cloud-based open-access artificial intelligence platform was constructed to provide assistance for detecting COVID-19 pneumonia worldwide. CT scan images could be uploaded freely by both clinicians and researches as an assistant tool, especially in other provinces or countries unfamiliar with the radiologic characteristics of COVID-19. This free open-access website can read images in batches, provide high-level auxiliary diagnostic services for different hospitals in free, and expand the boundaries of regions and manpower. Cases of COVID-19 pneumonia were also been made available on the open-access website, which might be a useful resource for radiologists and researchers for fighting COVID-19 pneumonia.

In summary, the deep learning-based model achieved a comparable performance with expert radiologist using much shorter time. It holds great potential to improve the efficiency of diagnosis, relieve the pressure of frontline radiologists, accelerates the diagnosis, isolation and treatment of COVID19 patients, and therefore contribute to the control of the epidemic.

## Data Availability

A website has been made available to provide free access to the present model. CT scan images could be uploaded by both clinicians and researches as a second opinion consulting service, especially in other provinces or countries unfamiliar with the radiologic characteristics of 2019-nCoV. Cases of 2019-nCoV pneumonia were also been made available on the open-access website, which might be a useful resource for radiologists and researchers for fighting 2019-nCoV pneumonia.

http://121.40.75.149/znyx-ncov/index

## AUTHORS’ CONTRIBUTIONS

YH and YL conceived and supervised the overall study. WL and GD contributed to writing of the manuscript. YH, YL, HS and WY contributed to critical revision of the report. CJ, ZL, ZJ, GD, WH, DZ, XY, ZY and CX contributed to collecting and analyzing the data of patients. CJ, ZL and ZY contributed to label CT images. HS, HX, ZB, ZK and WY developed the system. All authors reviewed and approved the final version of the manuscript.

## ACKNOWLEDGEMENTS

This work was partly supported by the grant from Novel Pneumonia Emergency Science and Technology Project of Hubei Provincial (to [Honggang Yu]), Hubei Province Major Science and Technology Innovation Project (Grant No. 2018-916-000-008 to [Honggang Yu]).

## FIGURES

**Supplementary Figure 1. The training curves of UNet++ for extracting valid areas in Computed tomography images**.

**Supplementary Figure 2. The training curves of UNet++ for detecting suspicious lesions in Computed tomography images**.

